# A standardised, high-throughput approach to diagnostic group testing method validation

**DOI:** 10.1101/2024.04.17.24305965

**Authors:** Leonhardt A. H. Unruh, Michael A. Crone, Paul S. Freemont, Leonid Chindelevitch

## Abstract

1

**Background:** Group testing, combining the samples of multiple patients into a single pool to be tested for infection, is an approach to increase throughput in clinical diagnostic and population testing by reducing the number of tests required. In order to further increase the throughput and accuracy of these approaches, mathematicians regularly devise novel combinatorial methods. However, although these novel methods are easily validated in silico, they are often never implemented in diagnostic laboratories because of the lack of clear and standardised pathways to clinical validation.

**Methods:** We develop a standardised analytical workflow that makes use of high-throughput automation and virus-like particle standards to validate theoretical group testing approaches. We then utilise specially developed virus-like particles for SARS-CoV-2, Influenza A, Influenza B, and Respiratory Syncytial Virus (RSV) to develop and validate a novel multiplex group testing approach based on simulated annealing and Bayesian optimization. Our approach improves the inference of positive samples in group testing, leveraging the quantitative nature of RT-qPCR test results.

**Results:** Our results show a higher positive predictive value of our novel approach for the inference of positive samples compared to the standard approach using binary test outcomes. In large-scale surveillance testing our method can greatly reduce the number of false positive identifications. Our in vitro findings show the viability of group testing for multiplexed testing of respiratory infections and demonstrate the potential of a novel inference method. Both innovations increase the number of people that can be tested with the available resources, which is particularly important in low-resource settings.

## 2 Introduction

During the SARS-CoV-2 pandemic many countries struggled to scale testing capacity in attempts to contain and control disease spread. In the United Kingdom, this meant that at the beginning of the pandemic, diagnostic testing capacity was so constrained that testing was limited to clinical settings. This precipitated disease spread within care homes and led to thousands of deaths ^1^. As resource constraints began to ease, testing was expanded as part of a community testing programme called Test, Trace, and Isolate (TTI). This programme greatly increased access to testing, but long turnaround times made it challenging to use for clinical decision-making.

One way to meet the need for increased clinical and community testing capacity is group testing, a technique dating back to 1943 when the statistician Robert Dorfman proposed it as a method to test soldiers for syphilis during WWII ^2^. Group testing works by pooling multiple samples together into groups and only testing each group as a whole, greatly reducing the number of tests needed. Group testing for a single pathogen is very useful during an outbreak, where the goal is containment of a disease which is of clinical or public health importance. During the SARS-CoV-2 pandemic several research groups have shown that they could achieve a high sensitivity with groups of up to 32 individual samples ^3,4,5^, making it practical to expand comprehensive testing to entire cities ^6^.

Group testing methods are typically classified as either adaptive or non-adaptive ^7^. Adaptive group testing allows the tests to be conducted in several rounds, with the groups tested in subsequent rounds informed by the results of earlier rounds. On the other hand, non-adaptive group testing is a time-saving variant in which all group tests are specified prior to testing. Both have been proposed as a way to maximise the use of scarce testing resources and accelerate the testing process, including during the SARS-CoV-2 pandemic ^8^. In our work we focus on non-adaptive group testing. Notably, the specific method we evaluate requires 65% fewer tests compared to individual testing. Non-adaptive testing also only requires half the time of adaptive group testing when positive samples are found, as no additional rounds of testing need to be conducted. Further, non-adaptive testing has the advantage over adaptive testing that logistics can be handled more easily, as there is no need of keeping track which samples to retest. This will likely reduce the number of human errors during the testing process.

The need to scale testing during the pandemic pushed many regulatory boundaries, with one US company, Quest Diagnostics, receiving clearance for applying an adaptive group testing design in the United States ^9^. However, although group testing brings many benefits, it is still rarely used as a part of business-as-usual diagnostic laboratory testing. Group testing is typically limited to screening blood transfusion products ^10^, as well as some Hepatitis C screening in diagnostic laboratories ^11^. Outside of the pandemic context ^12,13^, there is very limited regulatory guidance from national health bodies on exactly how to implement group testing in a clinical laboratory context.

There is an urgent need for regulatory reform around the validation of diagnostic tests that will allow for novel laboratory developed approaches to be utilised, but standardisation and harmonisation of validation techniques will be integral to these changes ^14^. During the pandemic we were involved with efforts to recommend technical and molecular standards that would be necessary to improve the accuracy and reliability of population testing ^15^. Our involvement in these efforts stemmed from our own experience using virus-like particles (VLPs) as molecular standards to validate clinical diagnostics for SARS-CoV-2 detection, where we showed that our VLPs were perfect proxies for clinical samples ^16^. Our developed standards were also utilised to determine the scientific, technical and logistical feasibility of adaptive group testing in a clinical laboratory setting ^17^.

With no clear framework for validation of group testing methods we decided to adapt and expand our already established molecular standards platform to develop a standardised, high-throughput approach towards this goal. We make use of precisely quantified VLPs, perfect surrogates for infectious viruses, along with an acoustic liquid handler, to test and optimise a singleplex (detecting only a single virus) group testing method. We then use our newly developed, standardised approach to evaluate non-adaptive group testing for use in multiplexed infectious disease testing. We simultaneously tested for SARS-CoV-2, Influenza A, Influenza B and RSV via multiplexed RT-qPCR. In conjunction with this validation we also developed `gtMAP` (available as R package), a novel computational approach to identify the patients infected with each of these viruses in group testing scenarios that makes use of the quantitative output of RT-qPCR, namely, the cycle threshold (Ct) value. `gtMAP` expands upon most standard group testing approaches which assume that tests are qualitative (with few exceptions ^18,19^), using a combination of simulated annealing and maximum a posteriori (MAP) optimisation of individual sample Ct value estimates to arrive at optimal estimates of sample positivity.

## 3 Results

### 3.1 Standardised, high-throughput method for group testing validation

It can be difficult to garner interest from clinical laboratory staff when trying to test a diagnostic device or method developed in an academic lab. This is typically because clinical laboratories operate under strict regulatory frameworks, and the use of patient samples, which are valuable, finite resources, often requires ethics approval. Our own experience suggests that a standardised method, developed and used during initial validation in an academic environment, is far more likely to generate the supporting data necessary to engage with clinical laboratory staff and justify further clinical validation ^16^.

Given the benefit that group testing methods could bring to clinical diagnostics, we sought to develop a standardised, high-throughput approach that could be used to validate such methods. Our goal was to integrate in silico design (Design) together with molecular standards and high-throughput laboratory automation (Build and Test) and finally analysis and optimisation (Learn) to incorporate Engineering Biology principles into diagnostics validation (Figure 1).

**Figure 1:**
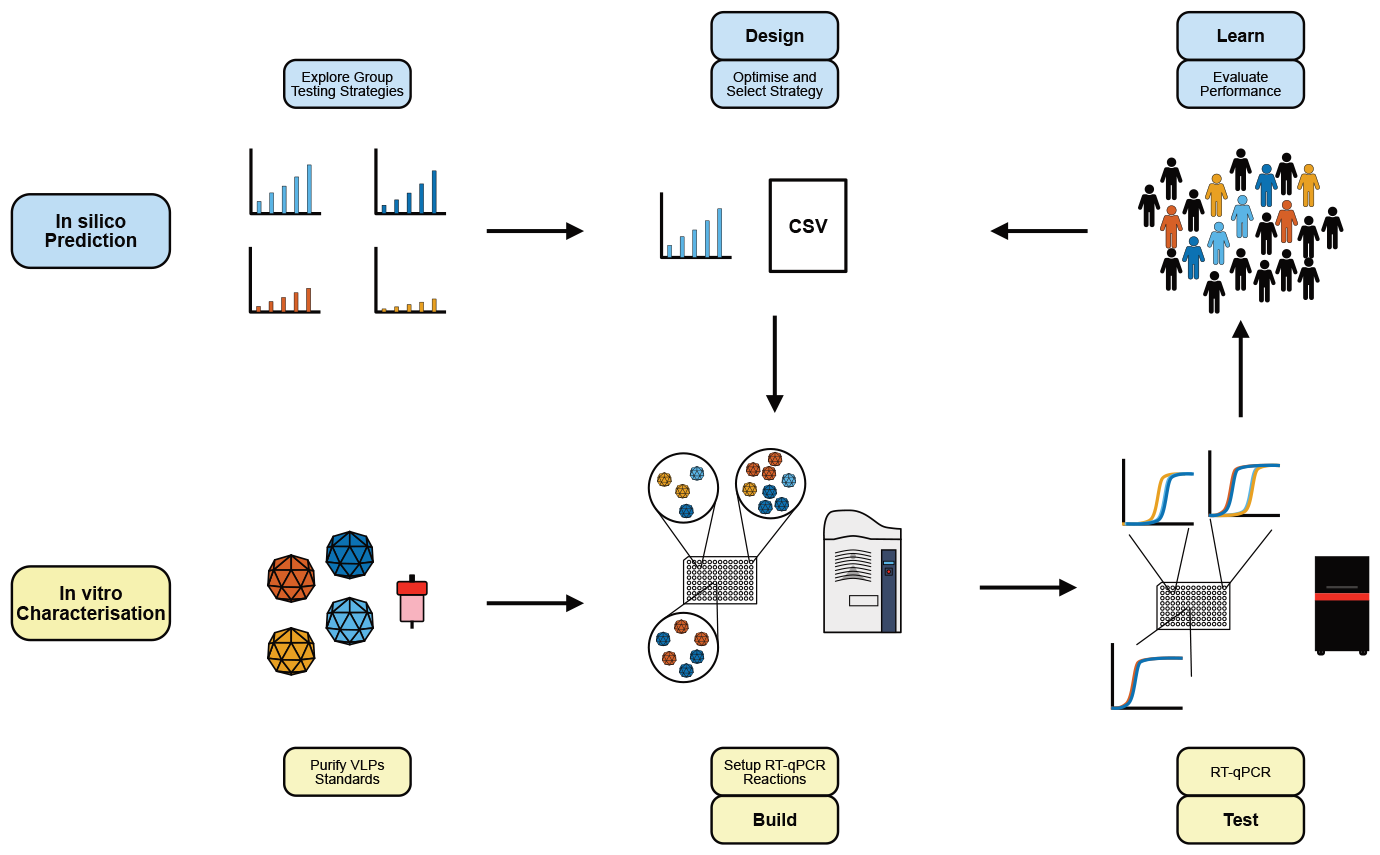
Schematic of the proposed standardised workflow for group testing validation. In silico methods are used to design testing strategies (Design). An acoustic liquid handler is then used to transfer virus-like particles and assemble RT-qPCR reactions (Build), which are subsequently analysed using RT-qPCR (Test). The results are then deconvoluted and the strategy optimised (Learn), which allows for further improvements with subsequent iterations.

Below we will first discuss our design and testing of an initial round of singleplexed testing for SARS-CoV-2, conducted as the first design step. Based on the promising initial results we then expand our method to multiplex testing. Next, we describe the development and characterisation of the VLPs used in the multiplexing experiments. Lastly, we present our results obtained from running a variety of multiplexing experiments and discuss the implication of these results for practical group testing as well as for the use of our validation procedure for future applications of group testing.

### 3.2 Singleplex group testing of SARS-CoV-2

We first performed the design stage on SARS-CoV-2 using a singleplex group testing strategy. As our first group testing design we chose a previously developed approach, the shifted transversal design ^20^ (STD), based on the concept of *d*-disjunctness. In a *d*-disjunct design, up to *d* positive items can be identified correctly without any error using binary (positive or negative) test outcomes. This design is chosen as it is easy to implement in practice and provides flexibility regarding the expected number of positive samples. Based on STD, we designed a decoding strategy for group testing implemented in the R package `gtMAP` and evaluated its performance in silico (see Supplementary Methods).

For our experiments we chose a 3-disjunct design with 81 samples tested in 36 groups containing 9 samples each. This choice allowed us to make efficient use of 96 wells, the standard plate format for most high-throughput RNA extraction and RT-qPCR machines. This design therefore optimizes for turnaround time by maximising the information obtained from a single setup.

For our singleplex build and test stages we used MS2 VLPs packaged with SARS-CoV-2 N-gene RNA suitable for detection with the CDC 2019-Novel Coronavirus RT-PCR Diagnostic Panel. We tested singleplex group testing with 3, 4, 5, and 6 positive samples out of 81, thus covering prevalence rates ranging from 3.7% to 7.4%. We placed the positive samples at random locations, with viral loads chosen uniformly at random from the interval [0-14000 copies/mL], ensuring that no group test had a final viral load of over 1700 copies/reaction in order to explore the lower limits of detection of our group testing strategy. We utilised 384-well plates, which are less commonly used for diagnostics, but more amenable for high-throughput in vitro characterisation of group testing. To keep with clinical practice we subdivided each 384 well plate into four 96 well plates. In each of these we placed 2 group testing setups, taking up 36 wells each. Each design was performed with 3 independent (biological) replicates that consisted of 3 technical replicates each.

For decoding of the PCR test results we compared both combinatorial orthogonal matching pursuit ^21^ (*COMP*), a standard binary decoder, and our novel decoder, `gtMAP` (illustrated in Figure 2), that makes use of the quantitative nature of PCR test results. `gtMAP` is based on a mixture of simulated annealing and MAP optimisation and was validated in silico (see Supplementary Methods). Briefly, `gtMAP` has two parameters that need to be determined by the user - the variance of Ct values and the positivity threshold. We set the variance of Ct values at 0.5 cycles. This value was chosen agnostically, rather than estimated from our laboratory results, to give a conservative estimate that would also apply to scenarios in which this variance cannot be reliably estimated. We set the decision threshold of the decoder to 0.5, based on calibration on simulated data. This setting means that a sample identified as positive in at least half the repeated decoder runs is declared positive, and otherwise it is declared negative (see Supplementary Methods for details).

**Figure 2:**
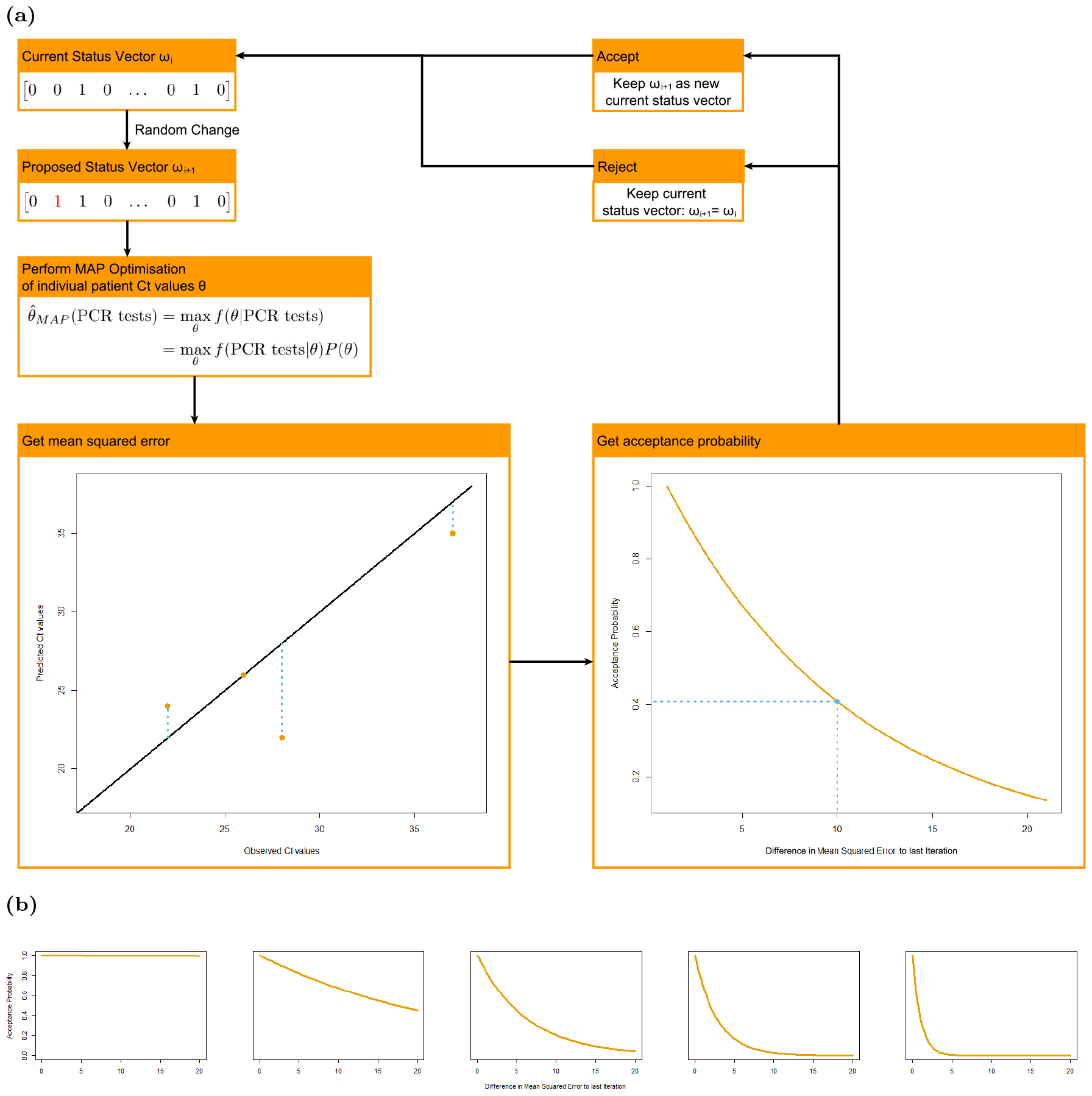
**(a)** Illustration of a decoding iteration. In each iteration *i* a status vector *ω*_*i*_ indicating positivity of the individual samples is randomly changed in one position from posititve (1) to negative (0), or vice versa. The resulting status vector *ω*_*i*+1_ is used as basis for a *maximum a posteriori* optimisation of the individual Ct values. Based on these individual Ct values, expected group test results are predicted and we obtain a mean squared error between these expected test results and the observed ones. Based on this mean squared errors’ difference to the mean squared error in the previous iteration as well as a temperature that declines with every iteration we determine the probability of accepting the proposed status vector *ω*_*i*+1_ or rejecting it and continuing with the current status vector *ω*_*i*_. The declining temperature at every iteration effectively causes the function of the acceptance probability to become steeper, such that we accept almost all proposals in the beginning, but are more strict in the end, accepting only improvements in the mean squared error (negative differences, for which the function is always equal to 1). The status vector after 500 iterations (with 10 iterations at each temperature) is then used to infer the positive samples. **(b)** Change in acceptance probabilities for different temperatures with temperature decreasing from left to right plots.

For the singleplex configurations tested, we observed the same performance for both `gtMAP` and the standard binary decoder. For 3 positive samples, the theoretical maximum number of positives samples that can be decoded without error for the design we chose, we recovered all the positives in each run without incurring any false positives, as expected. We showed that in silico, `gtMAP` generally outperformed the binary decoder with higher numbers of positives (see Supplementary Methods), while the latter still exhibited high specificity in the singeplex settings we tested in vitro, with most runs returning only up to one false positive. This is likely due to random placement of the positive samples favourable to decoding using COMP. In summary, our results clearly show the in vitro feasibility of singleplex group testing, enabling us to expand our design to multiplex scenarios.

### 3.3 Development and characterisation of VLPs for respiratory diseases

Multiplexed group testing is most effective when utilized for screening patients exhibiting symptoms potentially caused by various infectious agents. One such application is the testing for seasonal respiratory pathogens such as RSV and Influenza. Diagnosis is traditionally supported by the use of respiratory panels and the CDC have previously published a multiplex assay that detects Influenza A, Influenza B and SARS-CoV-2 ^22^ as well as an assay for the detection of RSV ^23^. However, these multiplex assays are typically used on single patient samples and multiplex assays for group testing have only been described for the diagnosis of Sexually Transmitted Infections ^24^. We first developed suitable molecular standards for the typical respiratory pathogens and then utilised them to validate our multiplex group testing approach.

First, virus-like particles (VLPs) were produced for each of the targets of the CDC multiplex assay and the RSV assay ^23^. All the viruses are RNA viruses and we could therefore make use of the VLP platform that we previously developed and utilised for clinical SARS-CoV-2 diagnostic workflow validation ^16^ (Figure 3). The Influenza A VLP packaged the Matrix CDS (Coding Sequence) from the A/Illinois/20/2018 (H1N1) strain. The Influenza B VLP packaged the Nuclear Export CDS from the B/Colorado/06/2017 strain. Our previously developed SARS-CoV-2 VLP needed to be adapted to package a different part of the SARS-CoV-2 genome and so a new VLP was produced that packaged the E, N and ORF10 CDS of the CoV USA/WA-CDC-WA1/2020 strain. The RSV VLP packaged the P and M CDS of GenBank accession M74568.1.

**Figure 3:**
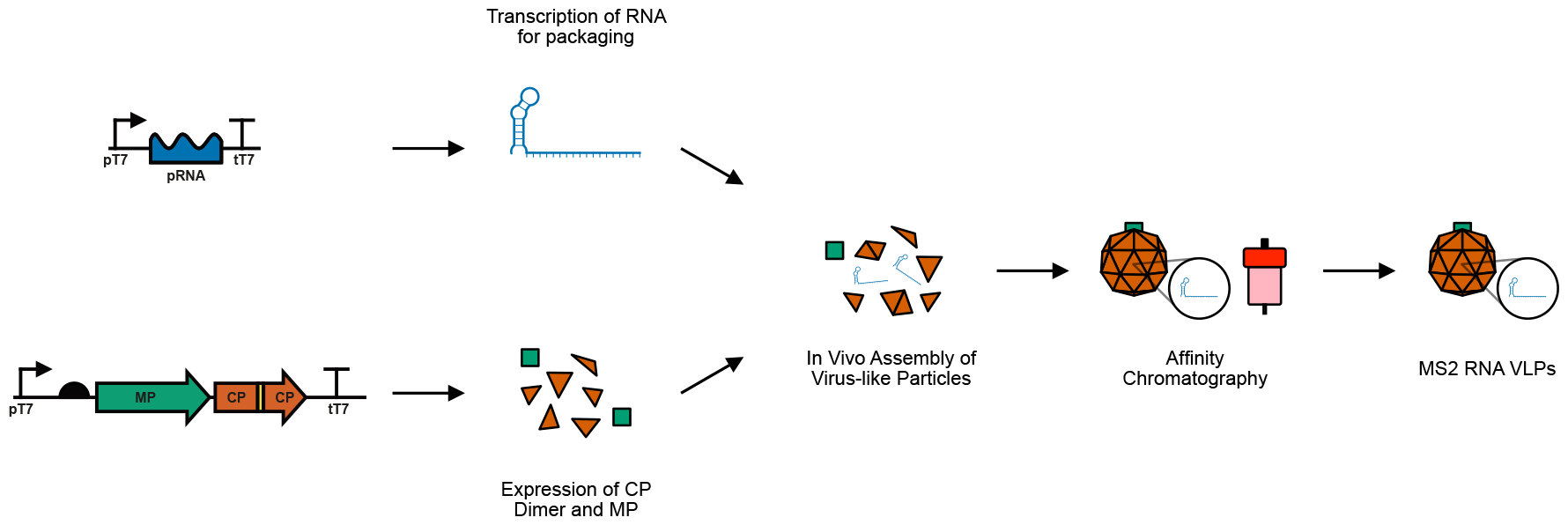
Workflow describing the steps in the production of MS2 RNA VLPs for various respiratory pathogens. The RNA to be packaged (pRNA) is transcribed while the maturation protein (MP) and coat protein (CP) dimer are expressed. Spontaneous assembly then occurs in vivo and the assembled particles are purified using affinity chromatography.

All VLPs underwent the standard biochemical and molecular characterisation performed previously for our SARS-CoV-2 VLP. This included RT-ddPCR to precisely quantify the VLPs and qPCR with and without reverse transcriptase to determine the level of DNA contamination (Supplementary Table **??** and Supplementary Figure **??**). This initial molecular characterisation also confirmed that the primer-probe sets successfully targeted the packaged RNA within the VLPs.

The CDC Flu SC2 assay usually includes an RNase P internal control which serves as a sample adequacy control. However, in the context of group testing the use of this target does not make sense because it merely detects the presence of human RNA in the pooled sample, which doesn’t rely on all samples in the pool having been sampled correctly. It was therefore possible to replace this internal control with an additional target, in this case, that of another respiratory pathogen, RSV, which can be severe in young and elderly populations ^25^. Thereby we arrived at a multiplex assay that can detect four highly relevant viral respiratory pathogens.

Finally, an experiment to determine the lower limit of detection was conducted on our newly developed assay. A serial dilution from 1000 VLP copies/reaction down to 10 copies/reaction was performed for all four assay targets (RSV, Influenza A, Influenza B and SARS-CoV-2). All assays, except the Influenza A primer-probe set, detected targets down to at least 10 copies/reaction (Supplementary Figure **??**), the equivalent of a concentration of approximately 500 copies per mL (assuming an extraction volume of 200 *μ*L, 50 *μ*L elution volume with 100% recovery and 5 *μ*L of eluted sample being added to each RT-qPCR reaction). It is difficult to compare these values to the LoDs obtained in the CDC assay ^22^ because the titres of 50% infectious doses were used instead of copies per reaction. However, validation data from the CDC assay does show that the Influenza A assay has a LoD that is 50 times greater than that for SARS-CoV-2 ^22^. These limits of detection are appropriate for the use of group testing for all four diseases as the medians of their viral load distributions are 3 to 5 orders of magnitude greater than their LoD (Table 1).

**Table 1:** Mean Viral Loads for SARS-CoV-2, RSV and Influenza A and B.

| Pathogen | Median viral load (copies/mL) | Reference |
| --- | --- | --- |
| SARS-CoV-2 | $1 \times 10^{6.5}$ | 26 |
| Influenza A | $1 \times 10^{8.1-8.5}$ | 27 |
| Influenza B | $\sim 1 \times 10^6$ | 28 |
| RSV | $4.5 \times 10^7$ | 29 |

### 3.4 In vitro validation of multiplex testing

Building on our successful singleplex experiments we performed a validation of group testing in a multiplex context using the validated MS2 VLPs for SARS-CoV-2, RSV, Influenza A, and Influenza B. We chose the same design and decoders as for our singleplex testing and tested them for 3 to 6 positive samples. The positive samples for each of the diseases were chosen randomly and independently across diseases. Viral loads for each sample were once again uniformly sampled from the interval [0-14000 copies/mL] while ensuring that no group test had a final viral load over 1700 copies/reaction. In the singleplex experiments we observed test setups that by chance were favourable for the standard decoder, showing a low number of false positives. Following singleplex testing, as part of the learn stage of our validation workflow, we decided to evaluate the differences between COMP and `gtMAP` observed in silico in more detail. With that we also validate our approach for more complicated configurations. To achieve this, we ensured that at least some of the designs tested in vitro led to fewer false positive identifications than obtained using COMP when decoding with `gtMAP` in silico. Importantly, this also means that we might incur more false negatives using `gtMAP`.

Our results are shown in Figure 4 and clearly demonstrate the feasibility of group testing in a multiplexing framework. With 3 positives we observed no false positives or false negatives for either decoder, again confirming the feasibility of achieving the theoretical best error-free performance. We also compared `gtMAP` to COMP for more than 3 positive samples. The results show that `gtMAP` does not incur any more false negatives, while inferring fewer false positives (Figure 4). Note that we observed one false negative for both decoders due to a failed PCR-test for one of the group tests for 5 positive samples. This single false negative was at the lower limit of detection of the assay.

**Figure 4:**
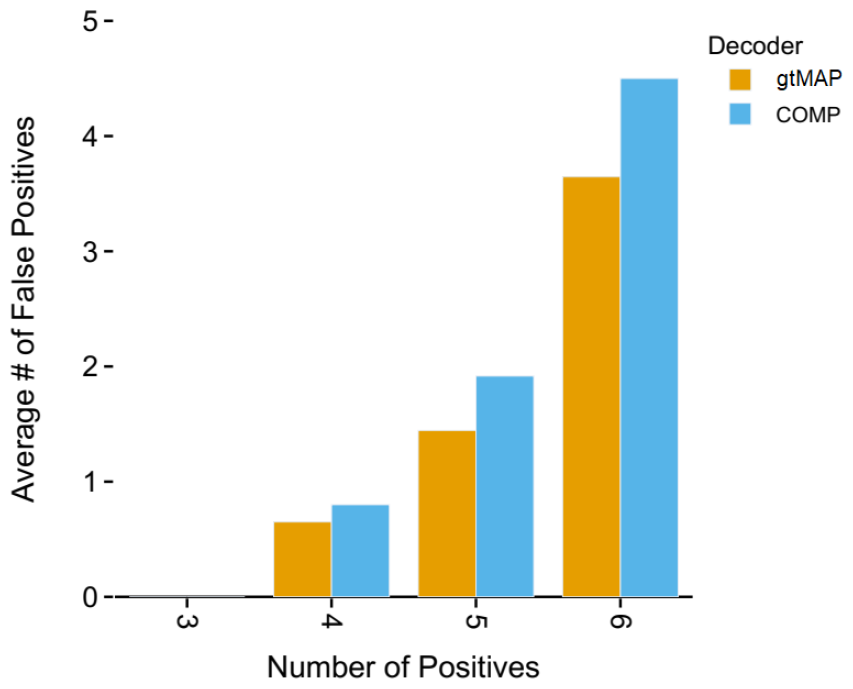
Average number of false positives in vitro for 3, 4, 5, and 6 positive samples out of 81 for `gtMAP` and COMP.

We further investigated the performance of `gtMAP` with varying decision thresholds (Figure 5). Our results indicate the suitability of the selected threshold of 0.5, as much higher thresholds lead to some false negatives. Overall, we showed that group testing is a viable strategy for multiplex PCR testing and that binary decoders can be improved upon when incorporating the quantitative information provided by PCR tests.

**Figure 5:**
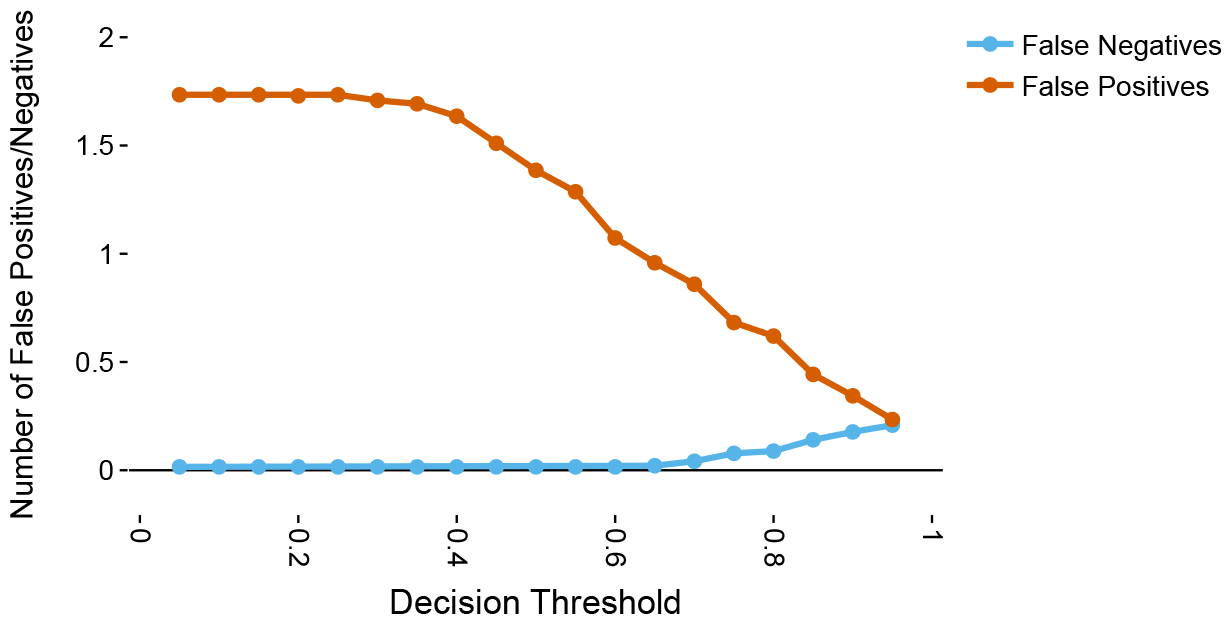
Average number of false positives (red) and false negatives (blue) across all number of positives for different decision thresholds.

## 4 Discussion

One of the greatest barriers to the implementation of novel group testing methods in clinical laboratories is the lack of a clear validation pathway in academic laboratories. During the COVID-19 pandemic we developed a standardised diagnostic validation pathway and utilised it to validate a novel, high-throughput, reagent agnostic RT-qPCR assay. Our developed assay was then used to perform almost 2 million tests in accredited clinical laboratories. In this work we describe how we have adapted this validation method to test the implementation of group testing. We start by showing that our validation method works for singleplex group testing. Based on these results, we subsequently develop and test a novel multiplex, non-adaptive group testing strategy. Our results show the high potential of applying group testing methodologies to multiplex respiratory disease testing, validating the methodologies’ application using a standardised workflow.

To the best of our knowledge, a thorough validation of non-adaptive group testing for multiplex testing has not been conducted previously. Our results show that group testing is a feasible approach in this setting, and that the chosen group testing design reliably achieves its theoretical capabilities. We chose a design that can identify up to 3 positive samples among 81 samples without error. This design saves over 55% of the tests relative to individual testing. Additionally, it requires less time to conduct than either standard testing, due to the lower number of tests, and adaptive group testing, as this requires multiple rounds of testing. We demonstrate that its performance remains good even at twice the prevalence it was originally designed for. This design can be easily adapted to varying prevalences, reducing its capacity when the prevalence is low, or increasing it when it is high; expanded versions of this design can perfectly identify up to 8 positive samples while still using fewer tests than individual testing. Importantly, by showing the successful application of our validation method, we provide a framework for future research to build upon. Within this framework, novel group testing designs and decoders can be easily validated and compared to previous designs. This will enable efficient testing of proposed designs to select those most suitable for clinical practice.

We further demonstrate the effectiveness of `gtMAP`, a decoder that leverages the quantitative nature of PCR test results in regimes that are prone to error under the standard binary decoder, i.e. when the number of positive samples exceeds the guaranteed error-free capacity of the design. Of particular importance is the higher positive predictive value of `gtMAP` (Supplementary Figure **??**), especially advantageous in settings where many individuals are tested, and most of them are negative. In such a setting, a reduction of false positive identifications will have a significant effect. While other non-binary group testing decoders have been proposed in the wake of the SARS-CoV-2 pandemic ^18,19^, none of them have been validated in a multiplex setting.

Group testing will likely play a valuable role in future outbreaks and pandemics because of its ability to increase testing capacity without substantial extra costs or reduction in accuracy. Two potential applications are representative sampling surveillance approaches and genomic surveillance programmes. A representative sampling strategy was implemented by the Office for National Statistics in the United Kingdom in their Coronavirus (COVID-19) Infection Survey ^30^, with participants recruited from random representative households and a total of 150,000 individuals tested every fortnight (approximately 200 tests per 100,000 people per day). The annual cost of the programme, at approximately £390 million ($482 million), makes it an unaffordable approach and it is unlikely that similar programmes will be feasible in more resource-constrained settings.

Genomic surveillance programmes also rely heavily on both representative sampling ^31^ and adequate testing capacity, and there are currently stark disparities in testing rates between high and low income countries ^32^. Countries during the pandemic often circumvented the lack of representative samples by sequencing higher numbers of patients, but this approach is neither scalable nor feasible in resource-constrained settings. Group testing would increase testing capacity and allow for “smart” sampling of the population while preserving financial and staffing resources.

Similarly, multiplex testing will be valuable in testing populations that experience outbreaks of multiple infectious agents at the same time by enabling parallel testing for multiple diseases. Combining it with group testing further increases the number of individuals that can be tested using multiplex testing. The combination of the two approaches will greatly help in dealing with the increased pressures on health systems from the circulation of multiple respiratory viruses in the population.

## Data Availability

In vitro data are available online at https://github.com/lahunruh/gtMAP

## 5 Acknowledgements

This work is supported by the UK Dementia Research Institute which receives its funding from UK DRI Ltd, funded by the UK Medical Research Council, Alzheimer’s Society and Alzheimer’s Research UK. We also acknowledge funding from UKRI-EPSRC (EP/R014000/1) and Community Jameel. LU and LC acknowledge funding from the MRC Centre for Global Infectious Disease Analysis (reference MR/X020258/1), funded by the UK Medical Research Council (MRC). This UK funded award is carried out in the frame of the Global Health EDCTP3 Joint Undertaking.

## 6 Author Contributions

LU performed group testing design and in silico work. MAC performed experiments, including VLP preparation and all RT-qPCR experiments. LC and PF supervised the work.

## 7 Code & Data Availability

`gtMAP` is available as an R package from GitHub (https://github.com/lahunruh/gtMAP). The in vitro group testing results are also available there.

