## Supplementary Methods for "A standardised, high-throughput approach to diagnostic group testing method validation"

### 1 Supplementary Methods

#### 1.1 Problem Outline

##### 1.1.1 Group Testing Paradigm

In a group testing design we want to test $n$ samples, where each sample $X_j$ is either positive (1) or negative (0), by splitting them up and grouping them into $T$ tests. A group testing design is specified by a $T \times n$ design matrix $M \in \{0,1\}^{T\times n}$, where the entry $M_{i,j} = 1$ if the $i$'th test contains the sample $X_j$ and 0 otherwise. In classical group testing, the vector of binary test results $Y$ is then obtained as

$$Y_i = \bigvee_{j\in[1,n]} M_{i,j} \wedge X_j,$$

where $\vee$ and $\wedge$ are the Boolean OR and AND operators, respectively. Given the test results $Y$ and the design matrix $M$ one tries to infer which of the samples are positive. The approach used to do so is called the decoder. One popular decoder, COMP [1], identifies a sample as positive if and only if all the groups containing it test positive. When assuming perfect tests, this method correctly classifies all those items that are definite negatives, and recovers all positives. It may, however, introduce some false positives.

While both COMP and our own decoder, `gtMAP`, can be applied to any group testing design, we chose one particular design family to evaluate them on, namely, the $d$-disjunct shifted transversal designs [2] (STD). $d$-disjunctness is defined by the fact that the union of any $d$ columns (tests) of $M$ does not contain any other column. This implies that the COMP decoder can correctly identify up to $d$ positive samples without error [2].

##### 1.1.2 PCR Test Results

PCR tests provide more than just positive or negative results. The outcome of one such test is the cycle threshold (Ct) value, which is proportional to the negative logarithm of the amount of virus, the viral load (VL), in the tested sample. Specifically, in a group testing design where the $j$-th sample has viral load $VL_j$ we can accurately model the group test results $Ct_i$ via

$$Ct_i = K - \log_2\left(\beta \sum_j \frac{M_{i,j} VL_j}{g_i}\right) + e, \tag{1}$$

where $K$ is a constant needed as Ct values are proportional to $-\log(VL)$, $\beta$ is a scaling factor, and $g_i$ is the number of items in the $i$-th group, called the group size. We assume that the measurement noise $e$ is drawn from a normal distribution, $e \sim \mathcal{N}(0, \epsilon)$. Intuitively, Equation 1 describes a relationship of Ct values and viral loads in which the Ct value of a group decreases from a baseline proportionally to the logarithm of the total viral load in the group. Importantly, the values for all of these parameters for different PCR-targets can be estimated from laboratory data. The novelty of our method lies in the use of these quantitative test results collected as a vector $\mathbf{Ct}$, instead of simply the binary result vector $Y$, to infer which items are positive.

#### 1.2 `gtMAP` decoder

To determine which samples are positive given $Ct$ and $M$ we employed a simulated annealing decoder combined with a maximum a posteriori (MAP) estimation approach. COMP is run prior to the simulated annealing step to reduce the problem size by excluding definite negative samples *a priori*.

**Simulated Annealing** The simulated annealing approach aims to estimate a binary status vector $\omega$ of which samples are positive (1) and negative (0). The status vector $\omega$ is initialized at random. At each iteration one sample is randomly changed from 0 to 1 or vice versa. The evaluation measure $E$ used is the sum of the squared errors of measured Ct values ($Ct_i$) and predicted Ct values ($PredCt_i$) given by a MAP optimisation of individual Ct values run with the current status vector,

$$E = \sum_i (Ct_i - PredCt_i)^2.$$

The acceptance probability is then determined by

$$P(accept) = \begin{cases} 1 & \text{if } E \leq 0 \\ e^{-E/T(\tau)} & \text{otherwise,} \end{cases}$$

where $T(\tau)$ is the current temperature. We established the best temperature decline function empirically as

$$T(\tau) = \frac{T_0}{1 + \tau^2 \alpha}$$

where $\tau$ is the current temperature iteration index (representing time) and $\alpha$ is a parameter set based on the inital temperature $T_0$. The starting temperature $T_0$ is determined by evaluating 10 random changes from the initialization vector $\omega$ over multiple iterations, at each either doubling or halving $T$ until the acceptance rate is between 0.7 and 0.9. $\alpha$ is then set such that in the final temperature iteration $T = 1$ (at $\tau_{max} = 50$). Overall, we iterated over 50 temperatures and had 10 iterations at each step. The 10 iterations at each step were chosen for computational speed and showed good results.

**MAP Optimisation** The MAP optimisation, implemented in RStan[3], aims to maximise the likelihood of the Ct values we predict, $PredCt_i$, given the measured group Ct values, i.e.

$$Ct_i \sim \mathcal{N}(PredCt, \epsilon^*)$$

where $\epsilon^*$ is our estimate for the standard deviation $\epsilon$ of a PCR-test results as in Equation (1). Our generative model underlying $PredCt_i$ is

$$PredCt_i = K^* - \log_2 \left( \beta^* \sum_j \frac{M_{i,j} 2^{IndCt_j}}{g} \right),$$

similar to Equation (1), but the viral loads are substituted by individual Ct values $IndCt_i$, i.e. the Ct values we would have observed had we tested the samples individually. We then place a normal prior on $IndCt_i$ to constrain them to a realistic range, i.e.

$$IndCt_i \sim \mathcal{N}(\mu, \sigma)$$

where we estimate the mean $\mu$ and the standard deviation $\sigma$ from individual test results, acquired from simulation. For clinical testing these would be obtained from previously conducted individual tests.

**Postprocessing** The initial decoding yields a set of candidate positives (samples $i$ with $\omega_i = 1$), but there is a risk of false negatives due to low viral loads. Specifically, a sample with a low viral load can be hidden by sharing all its tests with samples with a high viral load. These samples being inferred as negative (viral load of 0) or positive with a low viral load will only minimally impact the deviation of the inferred Ct values from the observed ones. Inspired by Wang et al. [4], we apply a postprocessing step by which we can identify the samples that could be hidden because they have a low viral load. To this end we iterate over the samples inferred as negative. One at a time, we set the individual Ct value of the current sample to a fixed minimum Ct value that we want to consider as potentially hidden, i.e. if the negative samples had this minimum individual Ct value, would their presence be detectable? Based on this we re-infer the predicted test values. Intuitively, a sample could be hidden if its viral load is low enough compared to the other samples it shares tests with such that the Ct value transform (Equation 1) of all the tests it is in would change by less than the assumed noise in Ct value generation. In this way, estimating the sample as positive would not change the Ct value estimates of the tests containing it more than expected by chance if it was negative. In practice, we infer that the sample in question could not be hidden even if it possessed a relatively low viral load if the absolute deviation between these new predicted test values and the originally inferred ones is larger than our estimate of $\sigma$. Otherwise, it could be hidden and we declare it positive.

##### 1.2.1 Decision Threshold

Running the MAP optimisation for every possible status vector of a group testing problem shows that there are multiple status vectors that provide similar results that would all be acceptable solutions under a given noise assumption for PCR tests. Additionally, one can also easily construct ambiguous combinations of positive samples giving two solutions equal validity (under a uniform prior assumption). Further, the simulated annealing algorithm has a random component. Hence, it is not surprising that repeated runs of the decoder return different optimal status vectors. However, this provides an opportunity for improved decoding. We assumed that some samples are more likely to be declared positive and that the true positive samples would be those that are determined to be positive in a large fraction of the solutions. Hence, by running the decoder multiple times (where we chose 20 repetitions) we can achieve a trade-off between sensitivity and specificity as mentioned in the main text by setting a decision threshold as the proportion of decoder runs above which we declare a sample positive.

##### 1.2.2 Generation of simulated Ct values

We first sampled VLs with an approach adapted from the model used in Cleary et al.[5]. Any individual will carry viral RNA for a certain amount of time, $t_{end}$. At first the virus will multiply exponentially within its host until a peak concentration is reached at $t_{max}$. After this the concentration will exponentially decline until $t_{end}$. VLs are then created according to

$$VL(t) = \begin{cases} 2^{\frac{\gamma}{t_{max}}t} & \text{if } 0 \leq t \leq t_{max} \\ \frac{2^{\gamma}}{2^{\frac{\gamma}{t_{end}-t_{max}}(t-t_{max})}} & \text{if } t_{max} < t \leq t_{end}, \end{cases}$$

where $2^{\gamma}$ is the maximum VL a host can carry. We chose $\gamma = 7.98$, $t_{end} = 17.62$, and $t_{max} = 3$ as in Cleary et al.[5]. For sampling of a single VL we pick the time $t$ uniformly at random from the interval $[0, t_{end}]$.

We transferred these back to Ct values plus noise via

$$Ct(VL) = Ct_{Baseline} - \log_2 \frac{VL}{VL_{Baseline}} + e,$$

where the noise $e$ was sampled from a normal distribution $e \sim \mathcal{N}(0, \epsilon)$. We used $\epsilon = 0.5$. We estimated $Ct_{Baseline} = 37$ and $VL_{Baseline} = 100$ approximately from the data provided in Vogels et al.[6]. An estimation from our in vitro validation data would also have been possible, however, estimating these parameters from a different source provides a more strict test of our methods' robustness.

##### 1.2.3 Optimization of `gtMAP`

By design, `gtMAP` requires the tuning of the temperature decline function, the prior on the individual Ct values, and the minimum Ct value for potentially hidden samples. To establish these we ran a thousand simulations for each of 3 to 8 positive samples among 121 samples in total in a 3-disjunct STD design. First, we established the optimal temperature decline function. We tested the temperature decline functions

$$T(\tau) = T_0 - \alpha\tau$$

as well as

$$T(\tau) = T_0 * \alpha^{\tau}$$

and

$$T(\tau) = \frac{T_0}{1 + \alpha\tau^2}$$

determining the latter one as the best decline function. $\alpha$ was chosen in each optimisation run, such that the final temperature $T(\tau_{max}) = 1$. We compared using a uniform and a normal prior for the individual Ct values, fitted to 10,000 simulated individual Ct values. We determined the

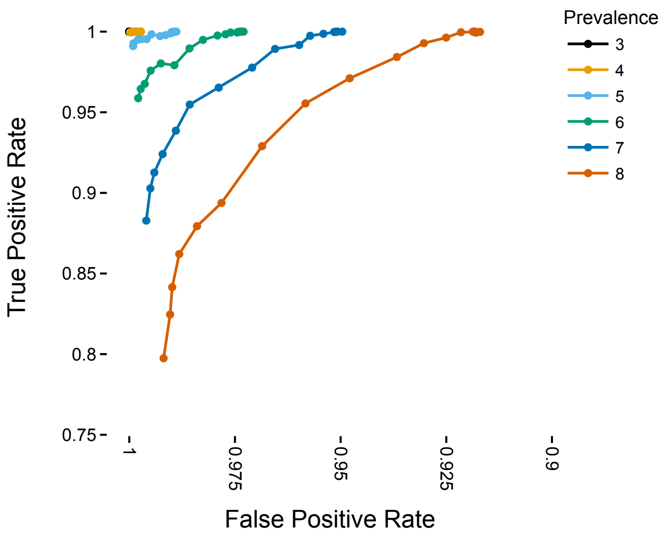


**Figure 1:** ROC curve for SA decoder at multiple thresholds for a true number of positives from 3 to 8 for $n = 81$ with a 3-disjunct STD design.

normal prior as the better prior. To establish the minimum Ct value we consider to be potentially hidden during post-processing we again ran 1000 simulations for the different number of positive samples. We compared values of 25, 30 and 35, and determined 25 to be the optimal value.

Further, we also tested choosing $K = 50$ and $\beta = 1$ for the Ct value estimation in the MAP optimisation, effectively allowing the viral loads to vary over a wider range and removing the scaling of viral loads. We observed better performance with these settings and chose them in our final decoder version. This also allows more flexibility in the case where $K$ is mis-estimated. In PCR-tests, Ct values usually do not go far above 40, hence $K = 50$ will likely always exceed the ground truth value, enabling estimation of all Ct values observed in the laboratory. Lastly, we tested different decision thresholds, running 20 iterations of the decoder and testing each threshold of positive sample identifications needed for a final declaration of a sample as positive between 0.05 and 0.95 in 0.05 increments (Figure 1). We picked 0.5 as the threshold that provided a good tradeoff between sensitivity and specificity.

#### 1.3 In silico validation

We tested our approach with a 3-disjunct STD design with 81 samples tested in 36 groups, containing 9 samples each. We compared our new method to COMP. First, we show that our new decoder is a generalization of COMP and additionally provides the option to fine-tune the expected specificity-sensitivity tradeoff. As group testing is an underspecified problem, there are generally multiple solutions to any configuration with more positives than the disjunctness of the design matrix (here, when $k > 3$). As discussed above, we set a decision threshold allowing us to obtain a specificity-sensitivity tradeoff shown in the ROC curves in Figure 1. A threshold of 0 declares all samples provided to our decoder (i.e. those declared positive by COMP) as positive, so setting the decision threshold to 0 reduces `gtMAP` to COMP.

Based on the optimization of `gtMAP`'s parameters we chose a decision threshold of 0.5 for our main simulation. While this threshold can be optimized for different numbers of positive samples, as shown in Figure 1, we found this value to be optimal for several settings, and this uniform setting allows us to make `gtMAP` agnostic to the exact prevalence. For the standard deviation of Ct values we chose 0.5. We ran 1000 simulations for each test setup with between 3 and 8 positive samples. These results are compared with COMP in Figure 2. We see that we achieve a higher balanced accuracy BA (defined as the mean of the specificity and the sensitivity, and thus prevalence-agnostic) than COMP, as shown in Figure 2F. We also observe a higher positive

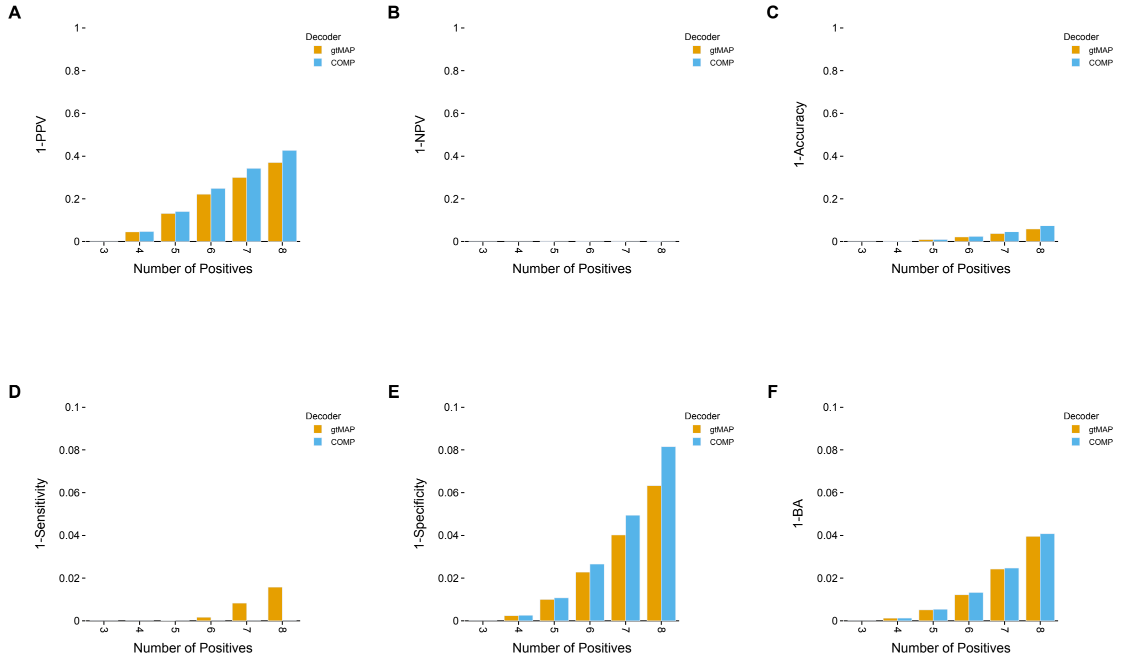


**Figure 2:** (A) PPV, (B) NPV, (C) accuracy, (D) sensitivity, (E) specificity, and (F) balanced accuracy (BA) of the COMP and `gtMAP` for 81 samples in a 3-disjunct STD design.

predictive value (PPV, Figure 2A), which is beneficial in a surveillance setting, as most people tested will be negative, increasing the overall number of correct predictions. We further report a higher overall accuracy of classification using our decoder (Figure 2C).

#### 1.4 Protein Purification of SARS-CoV-2 RNA Packaged MS2 VLP

The nucleic acid sequence of the N-gene of SARS-CoV-2 (accession number: NC_045512) was ordered from GeneArt (Thermo Fisher Scientific). The N-gene was cloned into a MS2 VLP expression plasmid backbone (Addgene #128233) using Type IIs assembly. The sequence-verified (Eurofins Genomics) plasmid (Addgene #155039) was transformed into Rosetta2™(DE3) pLysS cells (Merck). An overnight culture was used to inoculated 200 mL of Terrific Broth (Merck) supplemented with 50 mg/mL of Kanamycin (Merck), and grown at 30°C, 200 r.p.m. until an OD of 0.8. The culture was induced by supplementing with 0.5 mM IPTG (Merck) and grown at 30°C for a further 16 h. Cells were collected at 3220 × g at 4°C and stored at -20°C for later purification.

All protein purification steps were performed at 4°C. The cell pellet was resuspended in 4 mL Sonication Buffer (50 mM Tris-HCl pH 8.0, 5 mM $MgCl_2$, 5 mM $CaCl_2$, and 100 mM NaCl) with 700 U RNase A (Qiagen), 2500 U BaseMuncher Endonuclease (Abcam), and 200 U TURBO DNase (ThermoFisher Scientific). The cells were sonicated for a total of 2 min (50% amplitude, 30 s on, 30 s off) on wet ice. The lysate was then incubated for 3 h at 37°C. The lysate was centrifuged at 10,000 × g for 10 min at room temperature in a microcentrifuge. The supernatant was then filtered with a Minisart® 5 $\mu$m cellulose acetate (SFCA) filter (Sartorius) before being mixed 1:1 with 2× Binding Buffer (100 mM monosodium phosphate monohydrate pH 8.0, 30 mM Imidazole, 600 mM NaCl).

Supernatant was applied to a 5 mL HiTrap® TALON® Crude column (Cytiva) on an ÄKTA pure (Cytiva) primed with Binding Buffer (50 mM monosodium phosphate monohydrate pH 8.0, 15 mM Imidazole, 300 mM NaCl). The protein was eluted with elution buffer (50 mM monosodium phosphate monohydrate pH 8.0, 200 mM Imidazole, 300 mM NaCl) and then desalted and buffer exchanged into STE buffer (10 mM Tris-HCl pH 7.5, 1 mM EDTA, 100 mM NaCl) using an Amicon® Ultra-15 10 K Centrifuge Filter (Merck Millipore). The protein concentration was measured using the Qubit Protein Assay Kit and Qubit 3 Fluorometer (Thermo Fisher Scientific). The protein was then diluted in STE buffer, aliquoted, and stored at -80 °C.

#### 1.5 Reverse Transcription droplet digital PCR (RT-ddPCR) of SARS-CoV-2 VLPs

Reverse Transcriptase Droplet digital PCR was performed using the Bio-Rad QX200 Droplet Digital PCR system. Reactions were set up using the One-Step RT-ddPCR Advanced Kit for Probes (Bio-Rad) with primer and probe concentrations of 500 nM and 125 nM, respectively (primers and probes can be found in Table 1). The thermocycler settings used can be found in Table 2.

Data were exported in CSV format and analysed using a python implementation (`https://github.com/mcrone/plotlydefinerain`) of an online tool (`http://definetherain.org.uk`[7]). The online tool uses a positive control to define positive and negative droplets using K-means clustering, with rain being determined as anything outside three standard deviations from the mean of the positive and negative clusters. It then calculates final concentration based on

$$c = -\ln \frac{N_{neq}}{N} \frac{1}{V_{droplet}}$$

where

$$\begin{aligned} c &= \text{calculated concentration (copies/}\mu\text{L)} \\ N_{neg} &= \text{number of negative droplets} \\ N &= \text{total number of droplets} \\ V_{droplet} &= \text{average volume of each droplet } (0.91\ \times 10^{-3}\mu\text{L}) \end{aligned}$$

| Name | Forward Primer | Reverse Primer | Probe |
|---|---|---|---|
| 2019-nCoV_N1 | GAC CCC AAA ATC AGC GAA AT | TCT GGT TAC TGC CAG TTG AAT CTG | 5′-/FAM/ACC CCG CAT /ZEN/ TAC GTT TGG TGG ACC/3IABkFQ/-3′ |

**Table 1:** Primer and Probe Sequences for SARS-CoV-2 RT-ddPCR and RT-qPCR

| Stage | Temperature | Duration | Cycles |
|---|---|---|---|
| 1 | 50 °C | 60:00 | 1 |
| 2 | 95 °C | 10:00 | 1 |
| 3 | 95 °C<br>55 °C<br>72 °C | 0:30<br>1:00<br>1:00 | 50 |
| 4 | 98 °C | 10:00 | 1 |

**Table 2:** Thermocycler Settings for the RT-ddPCR for SARS-CoV-2 VLPs

#### 1.6 Reaction conditions for singleplex Group Testing

Primers, probes, and their relative concentrations were those recommended by the CDC and were ordered from IDT (primers and probes can be found in Table 1). TaqPath® 1-Step Multiplex Master Mix (Thermo Fisher Scientific) was used as the relevant master mix. RT-qPCR reactions were otherwise set up according to the manufacturer's instructions and thermocycling settings (according to the CDC protocol) except that total reaction volumes were 10 $\mu$L. A single mix with Master Mix, primers, probes and water up to 5 $\mu$L per reaction was added to each well in an Armadillo 384-Well plate (Thermo Scientific) using a multichannel pippette. Quantified SARS-CoV-2 MS2 VLP aliquots were extracted by heating to 95°C, diluted to 1000 copies/mL with Nuclease-Free Water and then transferred to an Echo® Qualified 384-Well Polypropylene 2.0 Plus Microplate. Additional wells with Nuclease-Free Water (Promega) were added which were then used to make up the remaining volumes for different reaction concentrations. In this way, the final 5 $\mu$L for each reaction was made up from the combination of varying quantities of VLPs and water. Liquid transfers were performed using an Echo 525 (Labcyte). Plates were sealed with MicroAmp Optical Adhesive Films (Thermo Fisher Scientific) and spun at 500 × g in a plate centrifuge. An Analytik Jena qTower3 84 was used for thermocycling according to the thermocycling settings in Table 3 and measurements were taken in the FAM channel.

| Stage | Temperature | Duration | Cycles |
|---|---|---|---|
| 1 | 25 °C | 2:00 | 1 |
| 2 | 50 °C | 15:00 | 1 |
| 3 | 95 °C | 2:00 | 1 |
| 4 | 95 °C<br>55 °C | 0:03<br>0:30 | 45 |

**Table 3:** Thermocycler Settings for Singleplex Group Testing

#### 1.7 Cloning and Expression of Respiratory Virus VLPs

The nucleic acid sequence of the N and ORF10 CDS of SARS-CoV-2 (strain: CoV USA/WA-CDC-WA1/2020), Matrix CDS of Influenza A (strain: A/Illinois/20/2018 (H1N1)), Nuclear Export CDS of Influenza B (strain: B/Colorado/06/2017) and P and M CDS of RSV (accession: M74568.1) were ordered from GeneArt (Thermo Fisher Scientific). The synthesised DNA for SARS-CoV-2, RSV and Influenza A were cloned into a MS2 VLP expression plasmid backbone (Addgene #128233) using Type IIs assembly with BsmBI. Due to an internal BsmBI site, the construct for the Influenza B was cloned by first digesting the Influenza B synthesised plasmid with BsaI and digesting the MS2 VLP expression plasmid backbone (Addgene #128233) with BsmBI. The two products were gel extracted and ligated with T4 ligase (Promega). The envelope CDS was added to the SARS-CoV-2 N and ORF10 CDS VLP using CPEC[8] with DpnI digested, gel extracted, PCR products generated using the primers in Table 4. The envelope CDS was PCR amplified from a previously generated plasmid pMC060-HisMS2_PLP_Env_pac (Addgene #155040).

The SARS-CoV-2, Influenza A, Influenza B and RSV VLP plasmids are available from Addgene under the openMTA. A summary can be found in Table 5.

| Template | Forward Primer | Reverse Primer |
|---|---|---|
| pMC060-HisMS2_PLP_Env_pac | TAC ACG GCC GCA TAA TCG AAA TTA ATA CGA CTC ACT ATA GGA TGT ACT CAT TCG TTT CGG AAG AGA CAG | CTG ATT TTG GGG TCC ATT ATC AGA CAT TTA GAC CAG AAG ATC AGG AAC TCT AG |
| HisMS2_VLP_SC2 | ATG TCT GAT AAT GGA CCC CAA AAT CAG | CTG TCT CTT CCG AAA CGA ATG AGT ACA TCC TAT AGT GAG TCG TAT TAA TTT CGA TTA TGC GGC CGT GTA |

**Table 4:** Primers Required for Cloning of Multiplex VLPs

| Plasmid Name | Pathogen | Gene | Strain | Addgene Plasmid # |
|---|---|---|---|---|
| HisMS2_VLP_FluA | Influenza A | Matrix CDS | A/Illinois/20/2018 (H1N1) | 175955 |
| HisMS2_VLP_FluB | Influenza B | Nuclear Export CDS | B/Colorado/06/2017 | 175956 |
| HisMS2_VLP_SC2 | SARS-CoV-2 | N and ORF10 CDS | CoV USA/WA-CDC-WA1/2020 | 175957 |
| HisMS2_VLP_EN_SC2 | SARS-CoV-2 | E, N and ORF10 CDS | CoV USA/WA-CDC-WA1/2020 | 175959 |
| HisMS2_VLP_RSV | Respiratory Syncytial | P and M CDS | GenBank: M74568.1 | 175958 |

**Table 5:** Summary of VLP plasmids for SARS-CoV-2, Influenza A, Influenza B, and RSV.

#### 1.8 DNA Contamination of VLP Preparations

Purified VLPs were analysed for DNA carryover during protein purification using qPCR with and without Reverse Transcriptase using the Luna Universal Probe One-Step RT-qPCR (NEB) kit. Reactions were setup with 1 $\mu$L of input VLP template that had been prepared by heating the sample for 95°C for 5 minutes. Primers and probes were as described in Table 6 and Table 7 and thermocycling settings can be found in Table 8.

| Name | Description | Oligonucleotide Sequence 5′ to 3′ | Concentration |
|---|---|---|---|
| InfA-F | InfA For1 | CAA GAC CAA TCY TGT CAC CTC TGA C | 3.33 $\mu$M |
| | InfA For2 | CAA GAC CAA TYC TGT CAC CTY TGA C | 3.33 $\mu$M |
| InfA-R | InfA Rev | GCA TTY TGG ACA AAV CGT CTA CG | 5.00 $\mu$M |
| | InfA Rev2 | GCA TTT TGG ATA AAG CGT CTA CG | 1.67 $\mu$M |
| InfB-F | InfB For | TCC TCA AYT CAC TCT TCG AGC G | 6.67 $\mu$M |
| InfB-R | InfB Rev | CGG TGC TCT TGA CCA AAT TGG | 6.67 $\mu$M |
| SC2-F | SC2 For | CTG CAG ATT TGG ATG ATT TCT CC | 6.67 $\mu$M |
| SC2-R | SC2 Rev | CCT TGT GTG GTC TGC ATG AGT TTA G | 6.67 $\mu$M |
| RSV-F | RSV For | GGC AAA TAT GGA AAC ATA CGT GAA | 6.67 $\mu$M |
| RSV-R | RSV Rev | TCT TTT TCT AGG ACA TTG TAY TGA ACA G | 6.67 $\mu$M |

**Table 6:** Flu SC2 Multiplex Assay Primers and Concentrations

| Name | Description | Oligonucleotide Sequence 5′ to 3′ | Concentration |
|---|---|---|---|
| InfA-P | InfA Probe | 5′-/FAM/TGC AGT CCT /ZEN/ CGC TCA CTG GGC ACG/3IABkFQ/-3′ | 1.67 $\mu$M |
| InfB-P | InfB Probe | 5′-/YakYel/CCA ATT CGA /ZEN/ GCA GCT GAA ACT GCG GTG/3IABkFQ/-3′ | 1.67 $\mu$M |
| SC2-P | SC2 Probe | 5′-/TexRd-XN/ATT GCA ACA /TAO/ ATC CAT GAG CAG TGC TGA CTC/3IAbRQSp/-3′ | 1.67 $\mu$M |
| RSV-P | RSV probe | 5′-/CY5/CTG TGT ATG /TAO/ TGG AGC CTT CGT GAA GCT/3IAbRQSp/-3′ | 1.67 $\mu$M |

**Table 7:** Flu SC2 Multiplex Assay Probes and Concentrations

| Stage | Temperature | Duration | Cycles |
|---|---|---|---|
| 1 | 25 °C | 2:00 | 1 |
| 2 | 50 °C | 15:00 | 1 |
| 3 | 95 °C | 2:00 | 1 |
| 4 | 95 °C<br>55 °C | 0:15<br>0:30 | 50 |

**Table 8:** Flu SC2 Multiplex Assay Thermocycler settings

#### 1.9 Determination of LOD of Multiplex Group Testing Assay

Reactions were setup using a Beckman Coulter Echo 525 with serial dilutions of VLPs from 1000 copies/reaction down to 10 copies per reaction of input VLP template that had been prepared by heating the sample for 95°C for 5 minutes. Primers and probes concentrations were as described in Table 6 and Table 7 and thermocycling settings can be found in Table 9. Reactions were setup according to the CDC's Influenza SARS-CoV-2 Multiplex Assay using the TaqPath™1-Step Multiplex Master Mix with 12.5 $\mu$L reaction volumes in 384-well Armadillo PCR plates (Thermo Fisher Scientific) sealed with ABsolute qPCR Plate Seals (Thermo Fisher Scientific) and thermocycled using an Analytik Jena qTower$^3$ 84. Reactions were set up in 96-well Armadillo PCR plates (Thermo Fisher Scientific) sealed with ABsolute qPCR Plate Seals (Thermo Fisher Scientific) and thermocycled using a Bio-Rad CFX96 Touch Real-Time PCR Detection System.

| Stage | Temperature | Duration | Cycles |
|---|---|---|---|
| 1 | 50 °C | 60:00 | 1 |
| 2 | 95 °C | 10:00 | 1 |
| 3 | 95 °C<br>55 °C | 0:30<br>1:00 | 40 |
| 4 | 98 °C | 10:00 | 1 |

**Table 9:** Thermocycler Settings for the RT-ddPCR for Multiplex VLPs

#### 1.10 Multiplex Group Testing Experiments

Primers, probes, and their relative concentrations were those recommended by the CDC and were ordered from IDT (primers and probes can be found in Table x). TaqPath® 1-Step Multiplex Master Mix (Thermo Fisher Scientic) was used as the relevant master mix. RT-qPCR reactions were otherwise set up according to the manufacturer's instructions and thermocycling settings (according to the CDC protocol) except that total reaction volumes were 12.5 $\mu$L. A single mix with Master Mix, primers, probes and water up to 7.5 $\mu$L per reaction was added to each well in an Armadillo 384-Well plate (Thermo Scientic) using a multichannel pippette. Quantified MS2 VLP aliquots of RSV, SARS-CoV-2, Influenza A and Influenza B were extracted by heating to 95°C, diluted to 2000 copies/$\mu$L with Nuclease-Free Water and then transferred to an Echo® Qualified 384-Well Polypropylene 2.0 Plus Microplate. Additional wells with Nuclease-Free Water (Promega) were added which were then used to make up the remaining volumes for different reaction concentrations. In this way, the final 5 $\mu$L for each reaction was made up from the combination of varying quantities of VLPs and water. Liquid transfers were performed using an Echo 525 (Labcyte). Plates were sealed with MicroAmp Optical Adhesive Films (Thermo Fisher Scientific) and spun at 500 $\times$ g in a plate centrifuge. An Analytik Jena qTower$^3$ 84 was used for thermocycling according to the thermocycling settings in Table 8 and measurements were taken in the FAM, YAK, TEX and Cy5 channels.

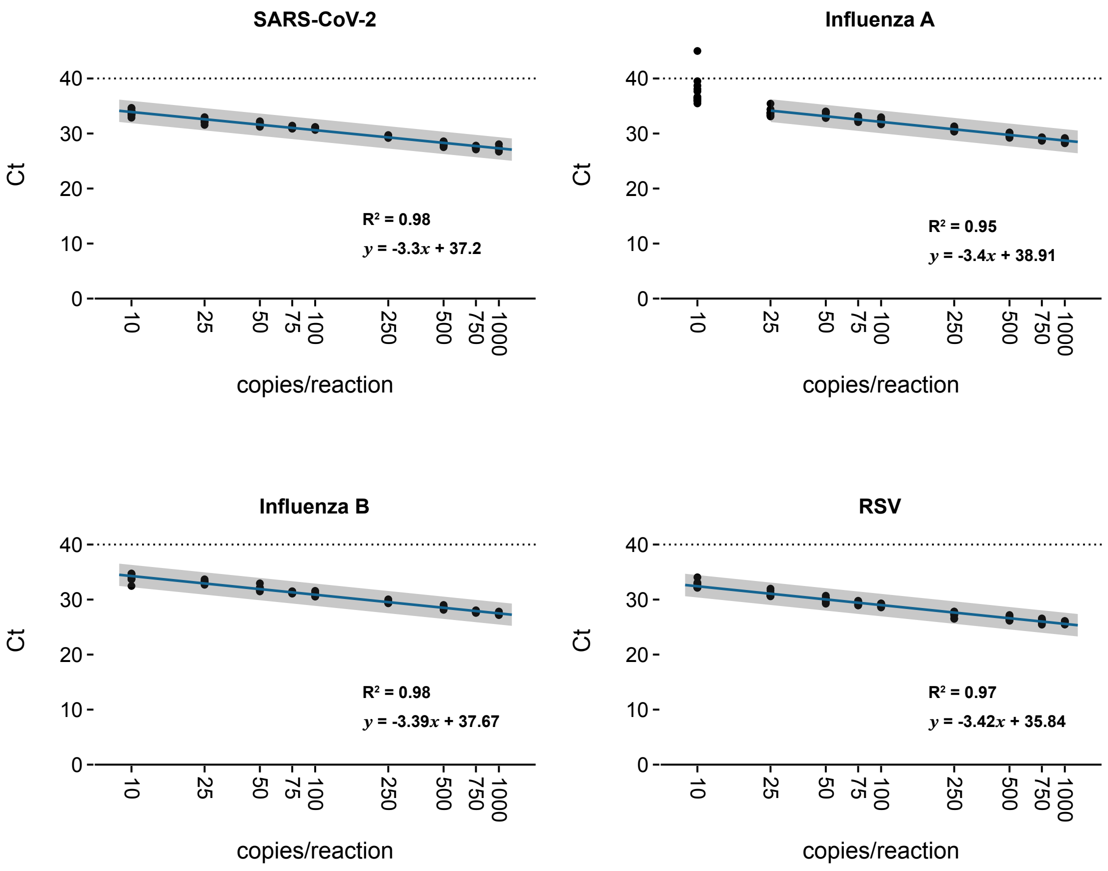


**Figure 3:** Multiplex assay for serially diluted VLPs from 1000 copies/reaction down to 10 copies/reaction showing detection down to at least 10 copies/reaction for all individual assay targets except Influenza A. Each serial dilution has 3 biological replicates made up of 4 technical replicates. The line is fit using linear regression with a 95% confidence interval shown in gray.

### 2 Supplementary Figures

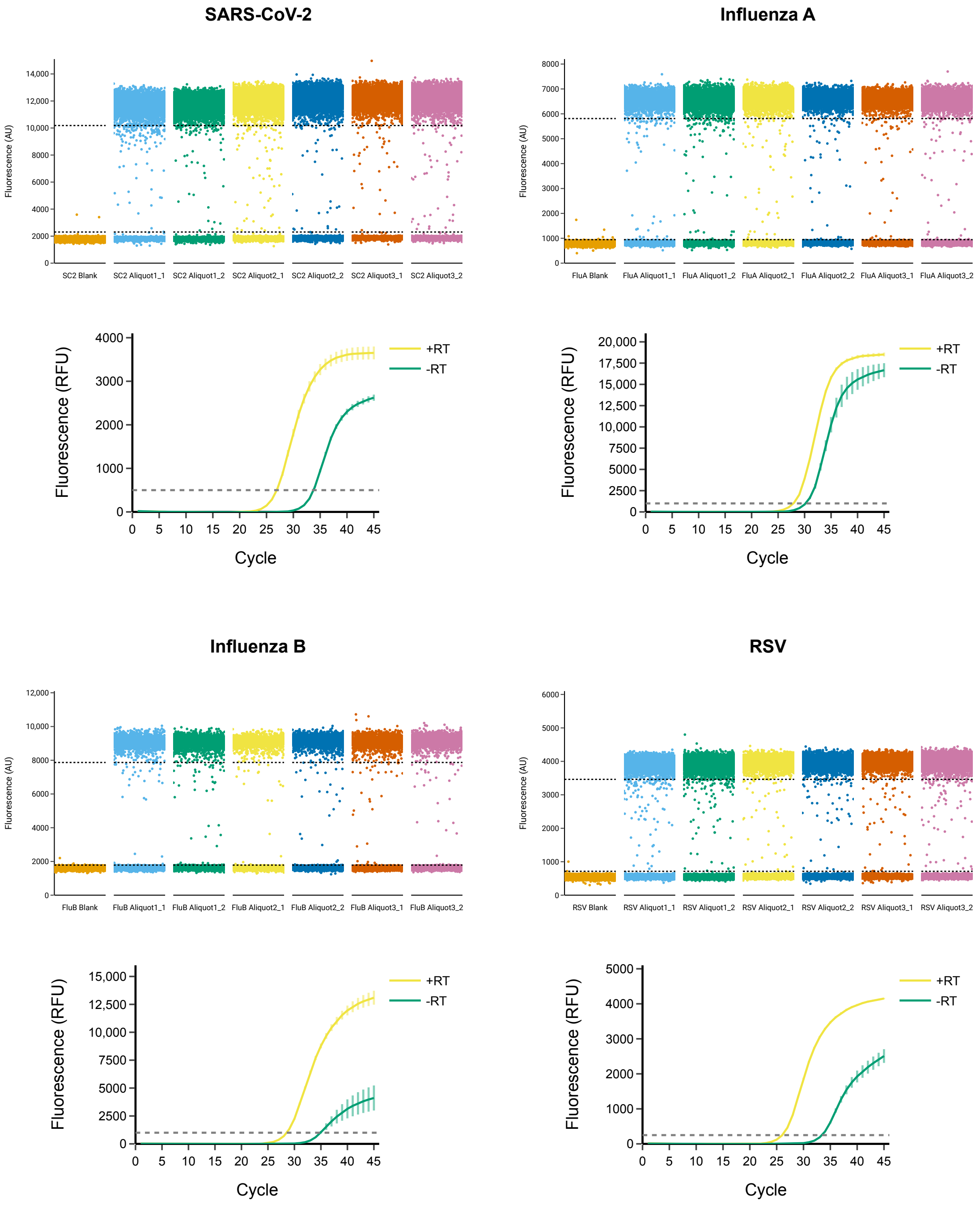


**Figure 4:** RT-ddPCR quantification of SARS-CoV-2, Influenza A, Influenza B and RSV VLP aliquots and PCR with and without Reverse Transcriptase (RT) to calculate the DNA contamination in each VLP preparation. RT-ddPCR was performed with two technical replicates and three biological replicates for each VLP. PCR was performed with 5 technical replicates.

**SARS-CoV-2**

| Aliquot | RT-ddPCR Positive Droplets | Mean | ± RT | qPCR Residual DNA Ct Value | Mean and SD |
|---|---|---|---|---|---|
| 1 | 22892 | 22788.5 | + | 27.16 | 27.05 |
| 1 | 22685 | | + | 27 | 0.089 |
| 2 | 23170 | 23184 | + | 27.01 | |
| 2 | 23198 | | − | 33.99 | 33.89 |
| 3 | 25325 | 25697 | − | 33.83 | 0.045 |
| 3 | 26069 | | − | 33.84 | |
| | Mean Copies | 23889.83 | | Ct Difference | 6.83 |
| | SD | 1577.50 | | % DNA | 0.88 |

**Influenza A**

| Aliquot | RT-ddPCR Positive Droplets | Mean | ± RT | qPCR Residual DNA Ct Value | Mean and SD |
|---|---|---|---|---|---|
| 1 | 7838 | 8059.5 | + | 27.34 | 27.33 |
| 1 | 8281 | | + | 27.31 | 0.016 |
| 2 | 9338 | 9548 | + | 27.34 | |
| 2 | 9758 | | − | 29.47 | 29.65 |
| 3 | 9936 | 9808.5 | − | 29.75 | 0.045 |
| 3 | 9681 | | − | 29.74 | |
| | Mean Copies | 9138.67 | | Ct Difference | 2.32 |
| | SD | 943.62 | | % DNA | 20.03 |

**Influenza B**

| Aliquot | RT-ddPCR Positive Droplets | Mean | ± RT | qPCR Residual DNA Ct Value | Mean and SD |
|---|---|---|---|---|---|
| 1 | 2432 | 2449 | + | 28.71 | 28.78 |
| 1 | 2466 | | + | 28.84 | 0.067 |
| 2 | 2259 | 2270.5 | + | 28.80 | |
| 2 | 2282 | | − | 34.99 | 35.32 |
| 3 | 2367 | 2310 | − | 35.32 | 0.045 |
| 3 | 2253 | | − | 35.65 | |
| | Mean Copies | 2343.17 | | Ct Difference | 6.54 |
| | SD | 93.76 | | % DNA | 1.08 |

**RSV**

| Aliquot | RT-ddPCR Positive Droplets | Mean | ± RT | qPCR Residual DNA Ct Value | Mean and SD |
|---|---|---|---|---|---|
| 1 | 14646 | 14632.5 | + | 25.91 | 25.87 |
| 1 | 14619 | | + | 25.85 | 0.031 |
| 2 | 13667 | 13801 | + | 25.86 | |
| 2 | 13935 | | − | 33.28 | 33.26 |
| 3 | 15178 | 15068.5 | − | 33.28 | 0.045 |
| 3 | 14959 | | − | 33.20 | |
| | Mean Copies | 14500.67 | | Ct Difference | 7.38 |
| | SD | 643.95 | | % DNA | 0.60 |

**Table 10:** Data from each produced VLP showing the absolute concentration per $\mu$L and the percent contaminating DNA for each preparation. Quantification was performed using RT-ddPCR with two technical replicates of three separate independent aliquots of the produced VLPs. Contaminating DNA was calculated by performing RT-qPCR with and without Reverse Transcriptase (RT). Contamination was calculated as a percentage of the total concentration.

[3] Stan Development Team. RStan: the R interface to Stan. URL `https://mc-stan.org/`. R package version 2.21.2.
